# Re-evaluating the melanoma TIL compartment and its unexpected spectrum of exhausted and functional T cells

**DOI:** 10.1101/2023.04.02.23288048

**Authors:** Cheryl M. Cameron, Brian Richardson, Jackelyn B. Golden, Yee Peng Phoon, Banumathi Tamilselvan, Lukas Pfannenstiel, Samjhana Thapaliya, Gustavo Roversi, Xing-Huang Gao, Leah L. Zagore, Mark J. Cameron, Brian R. Gastman

## Abstract

Significant heterogeneity exists within the tumor infiltrating CD8 T cell population, and exhausted T cells harbor a subpopulation that may be replicating and retain signatures of activation, with potential functional consequences in tumor progression. Dysfunctional immunity in the tumor microenvironment is associated with poor cancer outcomes, making exploration of these exhausted but activated (Tex/act) subpopulations critical to the improvement of therapeutic approaches. To investigate mechanisms associated with Tex/act cells, we sorted and performed transcriptional profiling of CD8^+^ tumor infiltrating lymphocytes (TIL) coexpressing the exhaustion markers PD-1 and TIM-3, from large volume melanoma tumors. We additionally performed immunologic phenotyping and functional validation, including at the single cell level, to identify potential mechanisms that underlie their dysfunctional phenotype. We identified novel dysregulated pathways in CD8^+^PD-1^+^TIM-3^+^ cells that have not been well studied in TIL; these include bile acid and peroxisome pathway-related metabolism, and mammalian target of rapamycin (mTOR) signaling pathways, which are highly correlated with immune checkpoint receptor expression. Through bioinformatic integration of immunophenotypic data and network analysis, we propose unexpected targets for therapies to rescue the immune response to tumors in melanoma.

## Introduction

Immune checkpoint inhibition (ICI) has revolutionized the treatment of many forms of cancer, especially UV induced skin cancers like melanoma ^1–6^. The success of ICI has also escalated the need for a comprehensive mechanistic understanding of the role cellular ICI targets play in oncogenesis and response to treatment. Furthermore, this successful clinical outcome has prompted a search for additional checkpoints and negative regulators of T cell function that can lead to new treatment options for the substantial subset of patients that are refractory ^1–3, 5–7^.

Although many immune cells in the tumor microenvironment (TME) express immune checkpoints like programmed cell death-1 (PD-1), it is the exhausted tumor infiltrating CD8^+^ T cell that was shown to be the key target of reinvigoration by ICI ^8–13^. Systems biology has revolutionized the comprehensive objective assessment of the state of T cells in myriad disease models. Transcriptional analysis via RNASeq has transformed the field of tumor immunology and facilitated the identification of new subpopulations within what was previously thought to be a homogeneous set of immune populations ^14, 15^. The vast majority of literature in melanoma, however, relies on small biopsies, so it is likely that the data may not represent the entire spectrum of alterations occurring in the immune system in the TME ^16, 17^. An integrated systems biology approach may yield key transcriptomic biological maps of the TME (and/or identify gaps) to provide insight into the delicate balance of regulation within the tumor and immune cells to pinpoint therapeutically targetable states within the spectrum of function.

We have a unique biobank of fresh, large volume tumors from surgical resections allowing us to sort down to very specific individual populations to investigate subset specific transcriptional regulation for discovery. We recently showed that the so-called “exhausted” CD8^+^PD-1^+^TIM-3^+^ TIL functionally inhibit healthy autologous T cell proliferation which can be inhibited by use of ICI therapies ^18, 19^. This finding contrasts with reports that these exhausted cells are dysfunctional, ^20, 21^. Interestingly, in the last few years several transcriptomic evaluations of CD8+ TIL identified exhausted T cell populations with high levels of activation ^22^ and even significant populations that are proliferating ^23^, however little has been done to investigate these TIL even though they express the same targets of modern ICI.

Given that we study actively suppressive CD8 T cells that express checkpoints, we hypothesized such cells are a subset amongst others generally considered to be exhausted T cells. We further hypothesized that investigation of CD8^+^PD-1^+^TIM-3^+^ T cells that have pathways indicating activation (Tex/act) will yield key transcriptomic information that can be exploited to identify new targets for immunotherapy. We strategically sorted and transcriptomically profiled CD8^+^PD-1^+^TIM-3^+^ TIL from melanoma and from cutaneous squamous cell carcinoma (SCC) tumors to answer these questions. We undertook what appears to be the largest unbiased transcriptomic evaluation (in terms of number of patients) of this key cellular ICI target within the TME and identified numerous differentially regulated biochemical pathways. Of note, these Tex/act TIL are metabolically active and demonstrate profound upregulation of pathways including bile acid metabolism, mammalian target of rapamycin (mTOR) signaling, and peroxisome-related pathways.

Given the fact that there is still a large group of melanoma and other cancer patients that do not derive benefit from ICI or other therapies, our findings may have broad application in helping reach the goal for therapeutic success in all patients.

## Results

### Transcriptomic profiling of dysfunctional CD8^+^ TIL

CD8^+^ TIL in melanoma and SCC share important common features, including the coordinate expression of immune checkpoint receptors and the development of resistance to immunotherapy ^18, 24, 25^, yet the mechanisms underlying any TIL dysfunction in these tumors remain elusive. To address this, we performed transcriptomic profiling on flow-sorted CD8^+^PD-1^+^TIM-3^+^ from melanoma and SCC (total CD8^+/^P^+/^T^+^ TIL) and compared them to peripheral CD8^+^ T cells (pCD8) from healthy donors. A total of 3,957 genes were differentially expressed (p≤0.05) in total CD8^+/^P^+/^T^+^ TIL vs. pCD8^+^ T cells. (Fig. 1A). Gene Set Variation Analysis (GSVA) beginning with gene ontology (GO) molecular function annotation identified the corresponding top upregulated pathways including chemokine activity, and three related to metabolism including carbohydrate kinase activity, oxidoreductase activity II, and aspartic type peptidase activity (Fig. 1A). The top downregulated pathways included C2H2 zinc finger domain binding, p53 binding, rRNA binding, ARF guanyl nucleotide exchange factory activity, and androgen receptor binding (Fig. 1A). Clear differential expression of genes in total CD8^+/^P^+/^T^+^ (SCC and MEL) vs. pCD8 indicates upregulation of immune checkpoint receptors including TIM-3 (HAVCR2), PD-1 (PDCD1), LAG3, and TIGIT (Fig. 1B).

**Figure 1:**
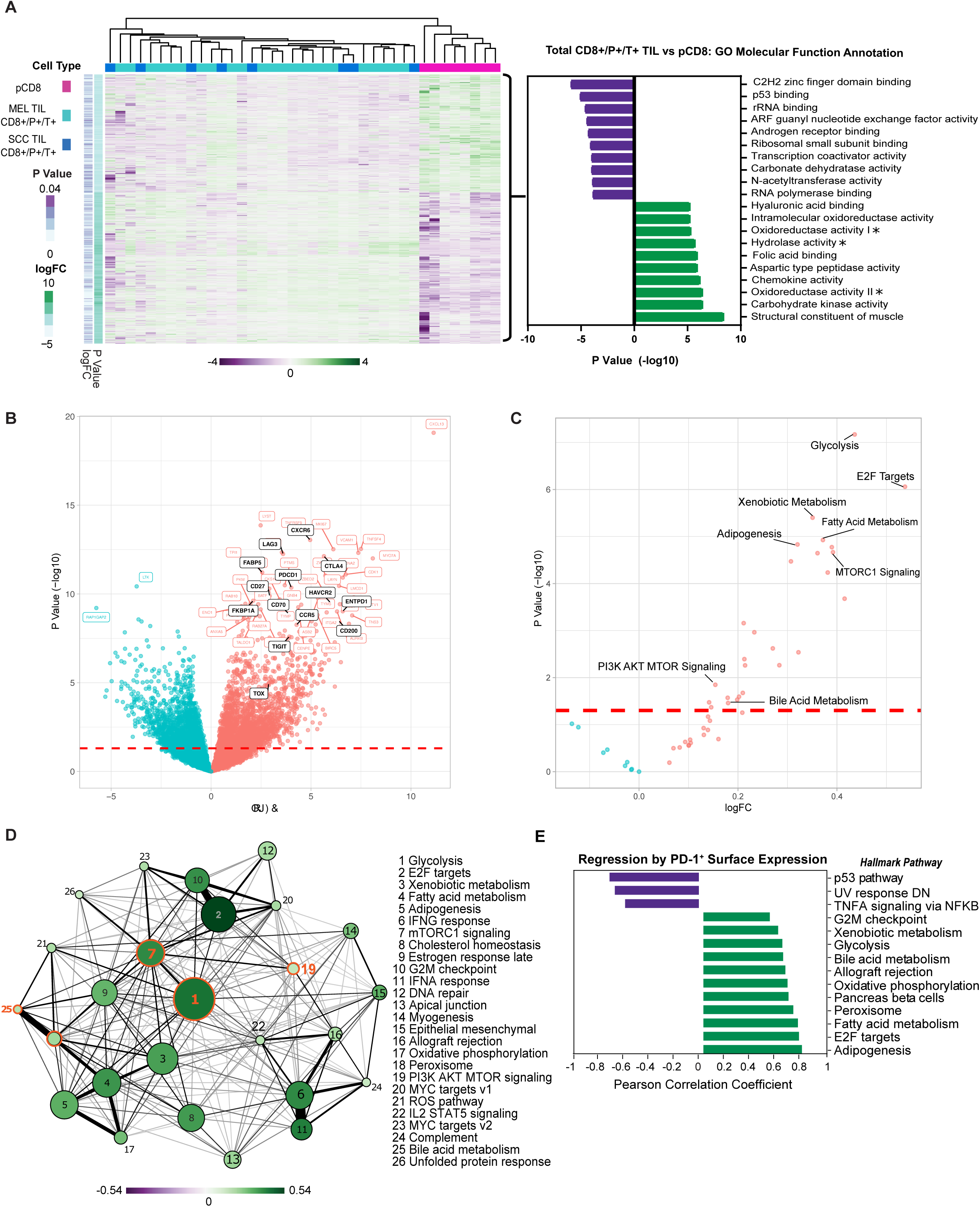
Melanoma and SCC TIL display a distinct transcriptional profile from CD8^+^ T cells in the periphery. **(A)** Heatmap of 3,957 differentially expressed genes (DEG) (nominal p<0.05) in PD-1^+^ TIM-3^+^ TIL from melanoma (MEL TIL CD8^+^/P^+^/T^+^) and SCC (SCC TIL CD8^+^/P^+^/T^+^) vs. pCD8 cells. Gene expression is normalized by z-score with green indicating higher relative levels of expression and purple indicating relatively lower levels of gene expression. GO Molecular Function analysis (via GSVA) of DGE in melanoma and SCC CD8^+^/P^+^/T^+^ TIL (total CD8^+^/P^+^/T^+^ TIL) vs. pCD8 is shown to the right of the heatmap. Asterisks indicate truncated pathway names; full pathways are as follows: Oxidoreductase activity acting on the CH-NH group of donors (oxidoreductase activity I), oxidoreductase activity acting on the CH-NH2 group of donors oxygen as acceptor (oxidoreductase activity II), and hydrolase activity acting on carbon nitrogen but not peptide bonds in cyclic amidines (hydrolase activity). **(B)** Volcano plot of the top 50 up- or downregulated genes in total CD8^+^/P^+^/T^+^ TIL vs. pCD8s. Upregulated genes are shown in red while downregulated genes are shown in blue, significant genes of interest to this study are annotated in black. The red dashed line indicates the significance cutoff (nominal p < 0.05). **(C)** Volcano plot of the Hallmark pathway enrichment in total CD8^+^/P^+^/T^+^ TIL vs. pCD8s. Significant pathways of interest are annotated; the red dashed line indicates the significance cutoff (nominal p < 0.05). **(D)** Enrichment map of significantly differentially enriched Hallmark pathways (nominal p <0.05) in total CD8^+^/P^+^/T^+^ TIL vs pCD8s. Node size indicates negative (-)log10(pvalue), node color with associated scale denotes log2 fold change values for enrichment scores while edge weight represents the Jaccard coefficient between significant sets of genes in each pathway. **(E)** Regression of PD-1 (GEOMEAN) surface expression on CD8^+^ T cells on sample pathway enrichment scores from the Hallmark database. Pathway enrichment scores are normalized by z-score with green indicating higher relative levels of enrichment and purple indicating relatively lower levels of enrichment.

Moreover, additional pathway enrichment analysis via GSEA using the Hallmark database (MSigDB) identified significant induction of multiple metabolic pathways in total CD8^+/^P^+/^T^+^ TIL including glycolysis, xenobiotic metabolism, fatty acid metabolism, mTORC1 signaling/PI3K AKT MTOR signaling, adipogenesis, and bile acid metabolism, corroborating our GSVA data (Fig. 1C). We performed analysis of pathway interactivity (via shared DEGs by Jaccard similarity coefficient) to provide a broader overview of how the enriched pathways interact, and as expected demonstrated a close correlation between mTORC1 signaling, PI3K AKT MTOR signaling, and glycolysis, with a less direct connection to bile acid metabolism (via mTORC1 signaling) (Fig. 1D). Glycolysis has been previously well described in TIL in melanoma and SCC; therefore, we were more intrigued with the significant differential enrichment of unexpected metabolic pathways, mTOR and bile acid metabolism in both cancers, which have not yet been proposed as key regulatory pathways in TIL function ^26^. Linear regression analysis of PD-1 (surface expression determined by flow cytometry) with gene expression revealed a strong positive correlation between PD-1 protein expression and upregulation of the bile acid metabolism, peroxisome, and glycolysis pathways in CD8^+^ TIL and PBMC (Fig. 1E).

Moreover, core biological pathways that have been previously shown to be associated with TGF-β attenuation of the tumor response to immune checkpoint blockade also were significantly differentially regulated within the dysfunctional CD8^+^PD-1^+^TIM-3^+^ melanoma and SCC TIL (Supp. Fig. 1A).

### Identification of unique metabolic-associated dysfunctional CD8^+^ T cell clusters by UMAP analysis

To integrate the expression of immune checkpoint receptors (ICRs), T cell phenotype, and activation markers with our transcriptional profiles, as well as to determine key pathways associated with TIL dysfunction, we carried out flow cytometric analysis of a randomly selected subset of MEL and SSC samples from tumors and peripheral blood (Fig. 2). We measured the expression of immune checkpoint receptors (PD-1, TIM-3, TIGIT, BTLA, LAG3, CD38) which have all been shown to regulate T cell function ^27^, with the hypothesis that coordinate expression of multiple immune checkpoint molecules will dramatically reduce T cell function.

**Figure 2:**
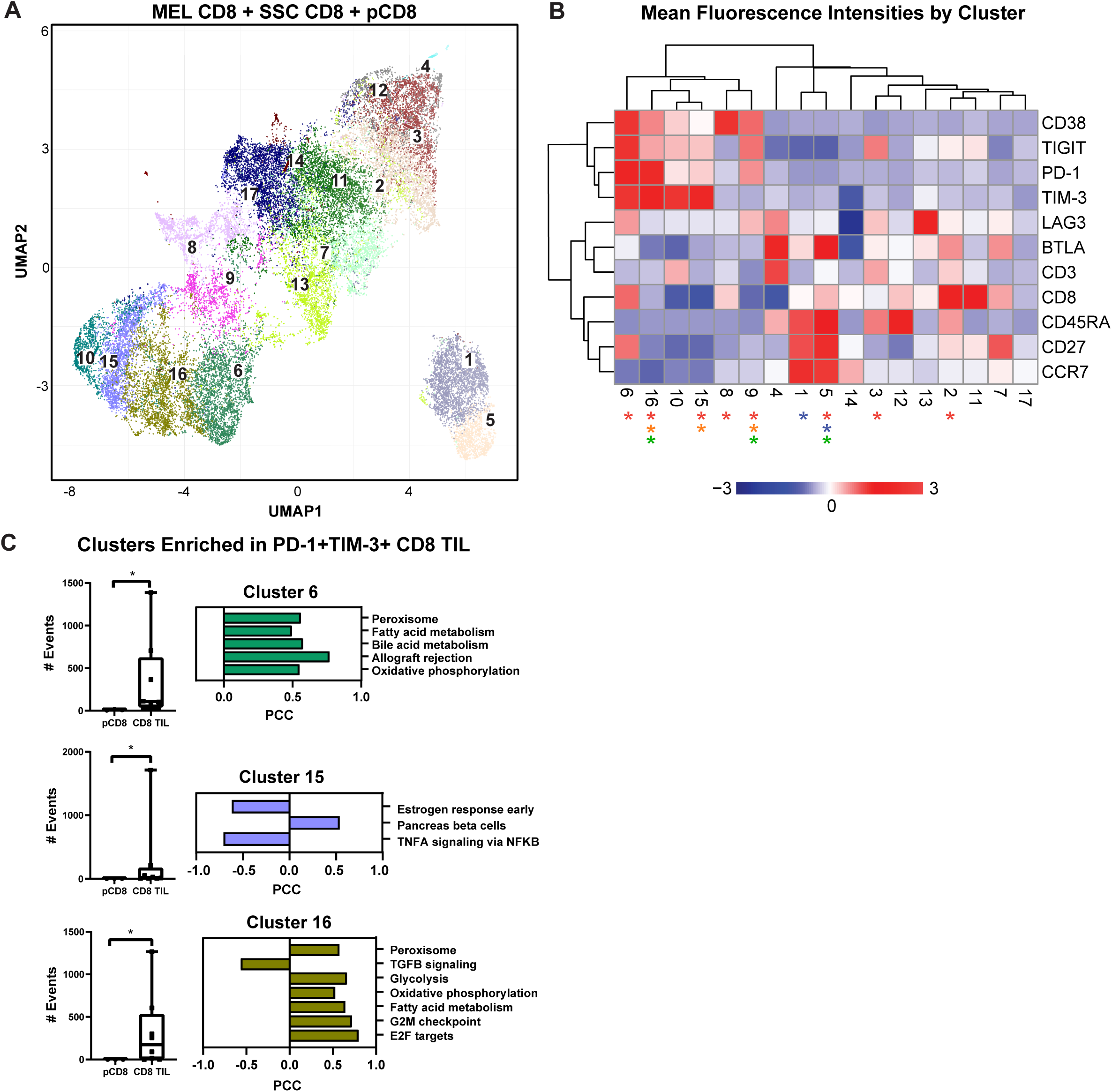
UMAP analysis and linear regression with immune checkpoint cell surface markers. **(A)** UMAP dimensional reduction analysis of CD8^+^ TIL compared to CD8^+^ PBMC reveals 17 unique clusters of coordinate cell surface protein expression. **(B)** Two-way hierarchically clustered heatmap of the MFI (geometric mean fluorescence intensity) of surface expression of CD38, TIGIT, PD-1, TIM-3, LAG3, BTLA, CD3, CD8, CD45RA, CD27, and CCR7 within the clusters in the UMAP analysis. Clusters significant for all TIL vs. pCD8 are indicated by red asterisks (2, 3, 5, 6, 8, 9, 15, 16), for MEL TIL vs. SCC TIL by blue asterisks (1, 5), for MEL TIL vs. pCD8 by orange asterisks (9, 15, 16), and SCC TIL vs. pCD8 by green asterisks (5, 9, 16). Heatmap is z-score transformed with upregulated MFI in red and downregulated MFI in blue. **(C)** Individual counts per selected cluster are shown in the box and whisker plots (clusters 6, 15, and 16 are notably enriched in all TIL compared to pCD8 cells; p<0.05; Mann-Whitney test). Linear regression modeling of each cluster with the RNAseq data reveals pathways enriched that are unique to each cluster.

We employed uniform manifold approximation and projection (UMAP), a nonlinear dimensionality-reduction machine learning approach, to analyze our multiparametric flow data and objectively determine the heterogeneity of CD8^+^ T cells based on surface expression of BTLA, CD3, CD8, CD27, CD38, CD45RA, CCR7, LAG3, PD-1, TIGIT, and TIM-3. UMAP analysis revealed 17 unique clusters, 9 of which show significantly different frequencies in CD8^+^ TIL compared to PBMC (Fig. 2A). A two-way hierarchically clustered heatmap of the MFI (geometric mean fluorescence intensity) based on surface expression showed the distinct surface signature profiles of the 17 clusters identified by UMAP analysis (Fig. 2B). Notably, the frequencies of lymphocytes co-expressing multiple immune checkpoints (CD38, TIGIT, LAG3, PD-1, TIM-3), the negative regulators of T cell function, were enriched in CD8^+^ TIL compared to periphery, especially in clusters 6, 15 and 16.

Of note, cluster 6 represents what is likely to be the most highly dysfunctional CD8 population, characterized by relative upregulation of CD38, TIGIT, PD-1, LAG3 and TIM-3, and is significantly enriched in the CD8^+^ TIL population compared to the peripheral CD8^+^ T cells (Fig. 2B and 2C). Similar to cluster 6, clusters 10, 15 and 16 have similar ICR expression patterns with increased coordinate immune checkpoint expression levels (Fig. 2B). Interestingly, cluster 16 is significantly enriched in CD8^+^ TIL of both melanoma and SCC with high expression of PD-1 and TIM-3 (Fig. 2B and 2C). Likewise, cluster 15 is also enriched in CD8^+^ TIL, but only in melanoma, with moderate immune checkpoint expression (Fig. 2B and 2C). Our findings indicate that clusters 6, 10, 15 and 16, with upregulation of ICRs, are likely to represent progressively more exhausted CD8^+^ T cell clusters (Fig. 2B).

We then performed linear regression modeling of gene expression with frequencies of cells from each flow UMAP cluster in each donor to identify transcriptomic pathway enrichment that correlates significantly with frequencies of CD8^+^ T cell clusters identified by UMAP. Interestingly, higher frequencies of cells in cluster 6 were strongly correlated with positive enrichment of the bile acid metabolism and peroxisome pathways (Fig. 2C). Likewise, cluster 16 frequency was also positively correlated with enrichment of the peroxisome and glycolysis pathways (Fig. 2C). In summary, our data demonstrate significant correlation between upregulation of immune checkpoint-expressing dysfunctional CD8^+^ TIL with activation of metabolic function, particularly bile acid metabolism, peroxidase and glycolysis pathways at the transcriptomic level. Targeting this metabolic-associated dysfunctional CD8^+^ Tex/act cell population could reinvigorate exhausted CD8^+^ T cells, thus enhancing immunotherapy responses in cancer patients.

### Upregulation of proliferation and metabolic associated genes in dysfunctional CD8^+^ TIL in melanoma

To determine whether these metabolic pathways were specifically dysregulated in melanoma, we performed differential gene expression analysis of the melanoma CD8^+^ TIL versus healthy peripheral CD8^+^ T cells resulting in a distinct gene expression signature as shown by the heatmap in Figure 3A. A total of 4,280 genes were differentially expressed in MEL CD8^+^ TIL versus peripheral CD8^+^ T cells, while 2,057 of those genes were commonly differentially regulated in MEL and SCC TIL (p≤0.05, Supp. Fig. 1B). The most statistically upregulated pathways using GO molecular function annotation (via GSVA) were oxidoreductase, structural constituent of muscle, and carbohydrate binding pathways (Fig. 3A). The top downregulated pathways include C2H2 zinc finger domain binding, N-acetyltransferase activity, rRNA binding, peptide N-acetyltransferase activity, and transcription coactivator activity. The top 50 differentially regulated genes between MEL TIL and pCD8 by statistical significance are shown in Figure 3B and the ICRs PD-1 (PDCD1), TIM-3 (HAVCR2), CTLA4 and LAG3 were among these. Also significantly upregulated were FABP5 (epidermal fatty acid binding protein) which can act as an intracellular receptor that binds to free fatty acids (FFA) in the cytosol and Ki-67 (MKI67), indicating enhanced proliferation of the CD8^+^ TIL.

**Figure 3:**
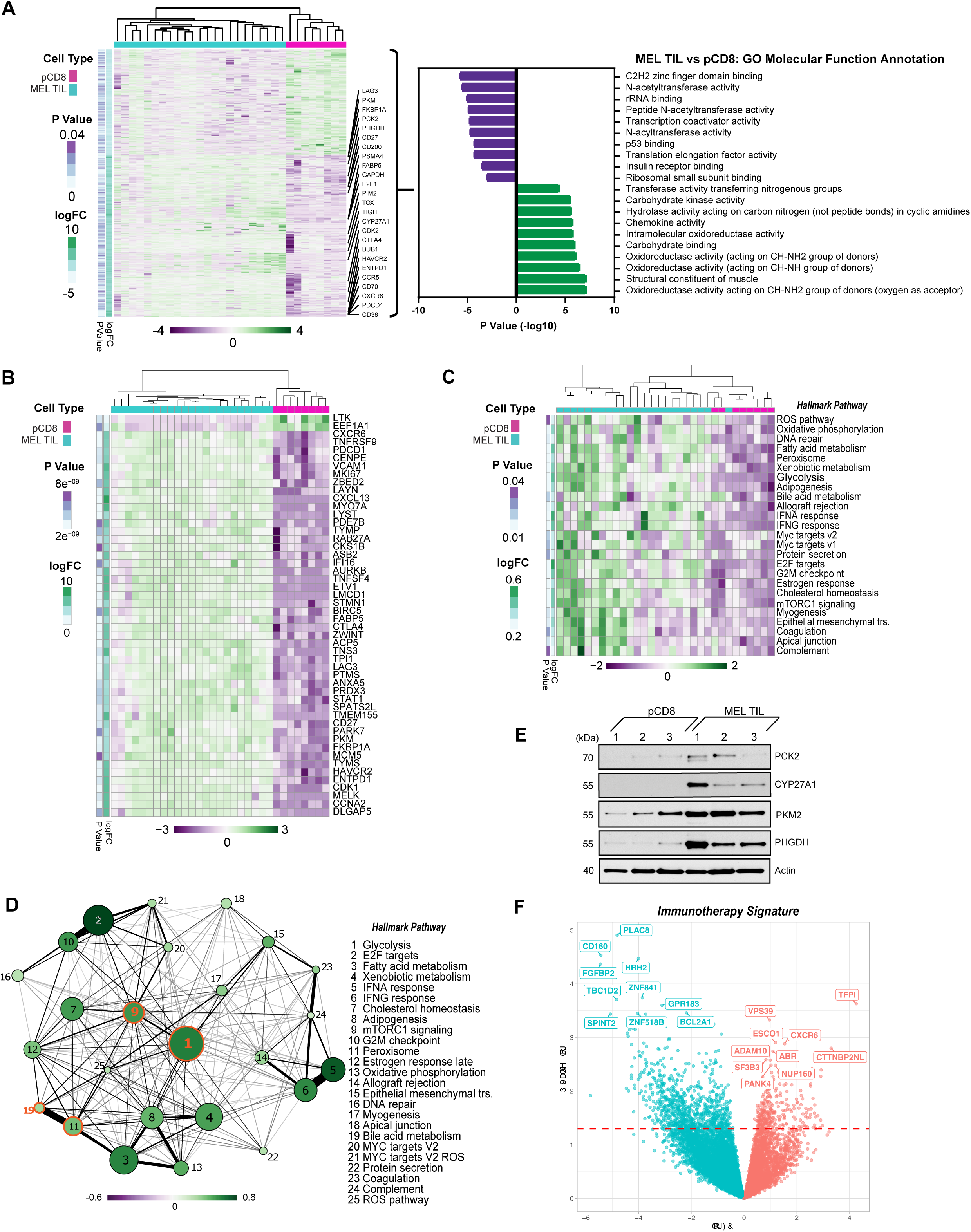
PD-1^+^TIM-3^+^ TIL from melanoma display a distinct transcriptional profile from CD8^+^ T cells in the periphery. **(A)** Heatmap of 4,280 differentially expressed genes (DEG) (nominal p<0.05) in TIL from melanoma vs. peripheral CD8 cells. Gene expression is normalized by z-score with green indicating higher relative levels of expression and purple indicating relatively lower levels of gene expression. Genes of interest are annotated. GO Molecular Function analysis (via GSVA) of DGE in melanoma TIL vs. pCD8 is shown to the right of the heatmap. **(B)** Top 50 differentially expressed genes from the MEL TIL vs. pCD8 contrast by p-value (nominal p < 8.93e-9). **(C)** Heatmap of significant (nominal p<0.05) Hallmark pathways associated with pCD8 versus MEL TIL. **(D)** Enrichment map of significantly differentially enriched Hallmark pathways (nominal p <0.05) in TIL from melanoma vs pCD8s. Node size indicates -log10(pvalue), node color with the associated scale denotes log2 fold change values for enrichment scores while edge weight represents the Jaccard coefficient between the sets of significant genes in each pathway. **(E)** Western blot analysis of control pCD8^+^ T cells (n=3) and melanoma TIL (n=3). Protein targets are shown on the right y-axis; β-actin was used as a loading control. **(F)** Volcano plot of 1072 differentially expressed genes (nominal p<0.05) in patients who have undergone immunotherapy (anti-PD-1). Upregulated genes are shown in red while downregulated genes are shown in blue; the top 10 up- and downregulated genes are annotated (red dashed line indicates p-value cutoff (nominal p<0.05)).

### Enrichment of metabolism pathways in CD8^+^ TIL in melanoma

To further evaluate the biology underlying differentially expressed genes found in CD8^+^ TIL and PBMC in melanoma, we performed pathway enrichment analysis of key canonical pathways. Notably, the peroxisome and bile acid metabolism pathways were coordinately regulated in melanoma TIL compared to pCD8 (Fig. 3C). Although a small number of reports have recently emerged showing the potential immunomodulatory effects of bile acids on the immune response, to the best of our knowledge ours is the first report of bile acid metabolism pathway dysregulation in tumor infiltrating lymphocytes in cancer ^28–30^. Not only are *peroxisome proliferator-activated receptor* (PPAR) pathways associated with FABP5, which was upregulated in dysfunctional CD8^+^ TIL, but they also regulate bile acid synthesis ^31–33^. Our data showed a strong positive enrichment of bile acid metabolism pathways in dysfunctional CD8^+^ T cells from melanoma tumors. One of the key regulatory enzymes of the bile acid metabolism pathway is CYP27, a cytochrome P450 enzyme encoded by the CYP27A1 gene ^34^. CYP27 metabolizes cholesterol into 27-hydroxycholesterol (27HC) through a hydroxylation reaction. Current evidence indicates 27HC can exert modulatory effects on the immune system and act as a selective estrogen receptor modulator (SERM) ^35, 36^. Strikingly, 27HC stimulates the proliferation of mouse melanoma cells by activating estrogen receptor alpha (ERα) and triggering the AKT and MAPK pathways ^37^. Compelling research into potential therapeutic strategies targeting bile acid metabolism can be found in breast cancer research ^38, 39^. For example, pharmacologic inhibition of CYP27A1 improved the efficacy of anti-PD-1 treatment and decreased metastatic breast cancer growth in mice ^40^. Additional studies have shown that CYP21A expression in human breast cancer tumors is correlated with disease progression ^38^. Notably, CYP27A1 was significantly upregulated in the CD8^+^ T cells of melanoma TIL as compared to PBMC (Fig. 3A). Upregulation of additional genes from the bile acid and peroxisome signaling pathways was also evident in TIL gene and pathway expression analysis, including SOD1, ACSL5, FADS2, CAT, DHCR24, SLC27A2, and IDH2 in melanoma TIL compared to pCD8s (Supp. Fig. 2).

As shown in Figure 3C, pathway enrichment analysis showed that the mTORC1 signaling pathway was significantly and positively enriched in CD8^+^ melanoma TIL as compared to pCD8s. The glycolysis pathway was highly integrated with many other pathways, particularly the mTORC1 and peroxisome pathways, which interact with bile acid metabolism (Fig. 3D). Compared to pCD8 T cells of control individuals, the glycolysis pathway was the most significantly upregulated pathway within the pathway interaction network analysis of the Hallmark canonical pathway database in CD8^+^ TIL of melanoma patients (Fig 3D). The differential regulation of multiple metabolic pathways, including bile acid metabolism, mTORC1, and glycolysis (Fig. 3D) likely reflects the fact that the CD8^+^ TIL cells are hosted in a hypoxic, acidic, and nutrient-depleted TME where the cellular metabolism has been reprogrammed as an adaptive mechanism to facilitate their proliferation and survival in melanoma. Our western blot analysis successfully revealed increased expression of CYP27A1, which was upregulated in the bile acid metabolic pathway, and three additional glycolytic genes (PKM2, PHGDH and PCK2) in melanoma CD8^+^ TIL compared to PBMCs (Fig. 3E).

### Identification of immunotherapy signatures

A subset of our cohort of melanoma patients were treated with anti-PD-1 immunotherapy. There were 1,072 differentially expressed genes (p≤0.05) in patients receiving therapy (n=5) as compared to those that did not receive treatment (n=8) as summarized in the volcano plot in Figure 3F. CXCR6 was upregulated in the melanoma anti-PD-1 immunotherapy signature, indicating a potential shift towards a proinflammatory tumor microenvironment which could lead to subsequent metastasis. CTTNBP2NL (CTTNBP2 N-terminal-like protein) was also upregulated and is known to be associated with STRIPAK complexes which have been broadly linked to metabolism, immune regulation, and cancer tumorigenesis ^41^. On the other hand, CD160 and HRH2 (histamine receptor H2) were downregulated in the anti-PD-1 therapy signature.

### mTOR Pathway Validation

The mTORC1 signaling pathway was highly significantly enriched in tumors as compared to peripheral blood (Figs. 1C and 3C). Using intracellular flow cytometry, we went further to discover that two of the key targets of mTORC1 complex activation, S6 ribosomal protein (target of S6 kinase which is a direct target of mTOR phosphorylation) and 4EBP1 (a direct target of mTOR phosphorylation), are significantly more phosphorylated in the MEL TIL compared to peripheral CD8 T cells, validating our findings at the phosphoprotein level (Fig. 4A). Furthermore, treatment with rapamycin (sirolimus), which specifically blocks MTORC1 signaling during TCR (anti-CD3/CD28) stimulation of melanoma TIL, resulted in consistent downregulation of PD-1 expression, particularly in the CD8^+^CD45RA^-^ memory T cells and CD8^+^CD45RA-CCR7^-^CD27^-^ effector memory T cells in the tumors (Fig. 4B).

**Figure 4:**
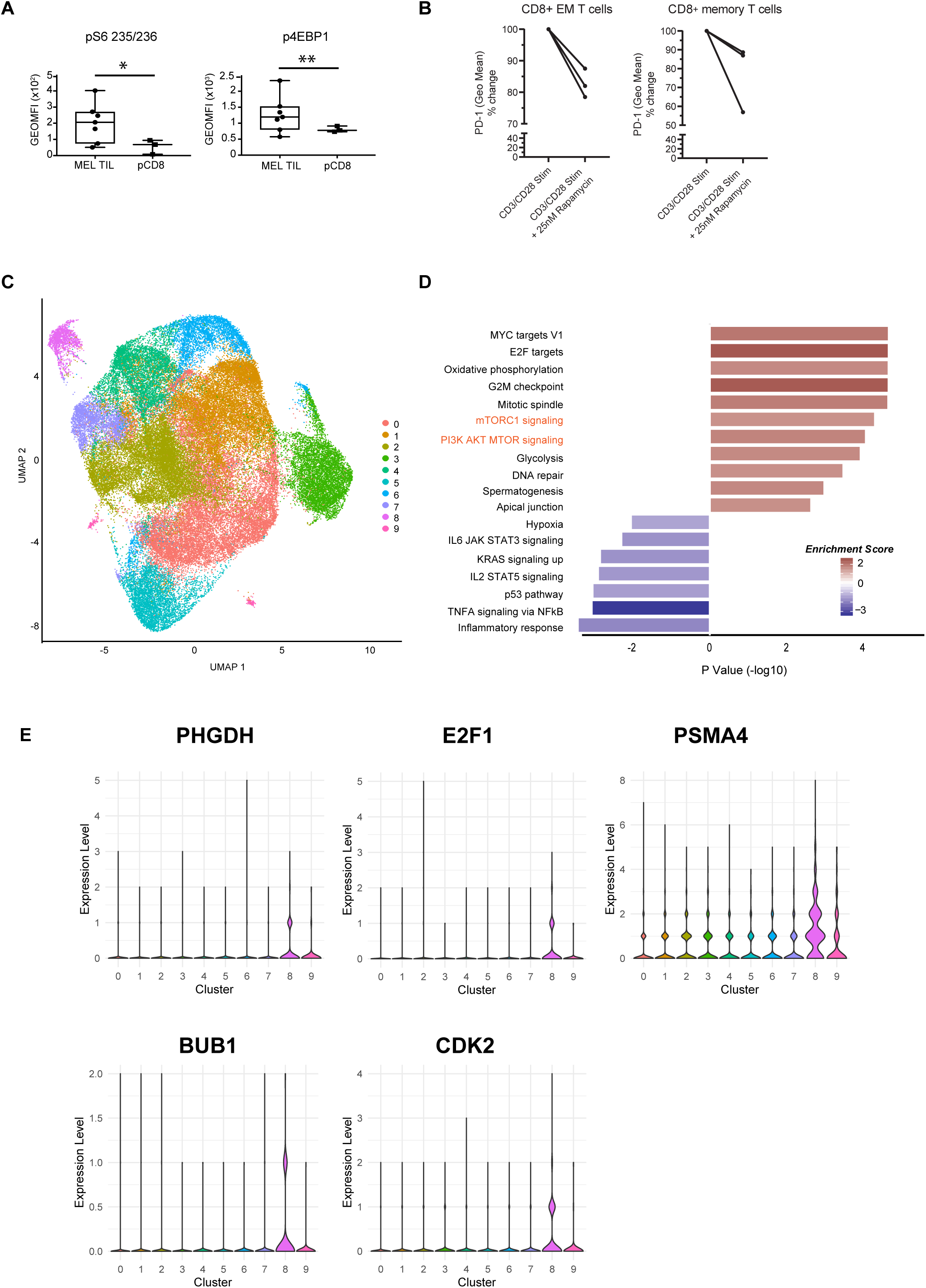
Validation of mTOR pathway (A) Intracellular flow cytometry of two phosphorylated targets, S6 ribosomal protein and 4EBP1, demonstrates significantly higher in the CD8^+^ TIL of melanoma compared to peripheral CD8^+^ T cells. *p=0.01 and **p=0.04 by Welch’s t-test **(B)** PD-1 expression is downregulated with rapamycin (25nM) treatment during CD3/CD28 stimulation (72hr) in melanoma TIL. CD8^+^CD45RA-CCR7^-^CD27^-^ effector memory T cells are shown on the left (*p=0.05, paired t-test) and CD8^+^CD45RA^-^ memory T cell trends are shown on the right (n.s.). **(C)** scRNAseq of CD8+ TIL in melanoma visualized with UMAP dimensional reduction analysis demonstrates 9 unique clusters. **(D)** Pathway analysis of Cluster 8 at single cell resolution demonstrates 18 significantly enriched pathways (nominal p<0.05) with positive enrichment of mTOR-related pathways such as mTORC1 and PI3K-AKT-MTOR (shown in orange text). **(E)** Violin plots of selected MTOR-related gene expression (PHGDH, E2F1, PSMA4, BUB1, CDK2) in the scRNAseq clusters.

Moreover, to validate differential enrichment of the mTOR pathway, we analyzed the single cell transcriptomes of CD8^+^ TIL from a new cohort of metastatic melanoma patients collected prior to anti-PD-1 immunotherapy. UMAP dimensional reduction of the single cell RNA sequencing (scRNAseq) dataset revealed a total of 9 clusters (Fig. 4C). Cluster 8 demonstrated positive enrichment of the mTORC1 and PI3K/AKT/MTOR signaling pathways at the single cell resolution, validating our bulkseq findings (Fig. 4D).

Violin plots of MTOR-related genes unique to cluster 8 (PHGDH, E2F1, PSMA4, BUB1, CDK2) are shown in Figure 4E, all of which were also differentially regulated in our bulk RNAseq signature of dysfunctional CD8^+^ TIL. We also confirmed PHGDH expression at the protein level (Fig. 3E).

## Discussion

Our study identified several novel gene signatures that distinguish CD8^+^ melanoma TIL from peripheral blood. More importantly, our findings reveal potential metabolic pathways that can be targeted to reinvigorate the dysfunctional immune system observed in melanoma and SCC-derived CD8^+^PD-1^+^TIM-3^+^ TIL. Through comprehensive transcriptomic analysis of immune checkpoint enriched CD8^+^ TIL and PBMC, we show that several metabolic signaling pathways, such as bile acid and peroxisome, along with mTOR pathways, are enriched in dysfunctional CD8^+^ Tex/act cells.

The observation of dysregulated bile acid metabolism in melanoma and SCC TIL is especially interesting given the considerable amount of data highlighting the relationship between antitumor immune response and the microbiome ^42–44^. The liver produces primary bile acids, which are then modified by gut microbes into a wide range of compounds with roles in gut metabolism, cell signaling, and microbe composition. Perturbations in the microbial ecosystem have been linked to immune evasion, carcinogenesis, and chronic inflammation and can affect the efficacy of cancer immunotherapies, including ICIs ^45^. While most studies have focused on the gut microbiome and its connection to immune surveillance dysregulation; the interplay between cancer cells, immune cells, and microbiota is also relevant in the tumor microenvironment ^46, 47^. Notably, it was observed that numerous tumor types, including melanoma, have distinct microbial signatures. The abundance of specific intratumor bacteria in melanoma has been linked to CD8^+^ T cell infiltration and patient survival ^48^. Additionally, patient immunotherapy response has been linked to metabolic functions encoded by a unique pattern of intratumor bacteria ^49^. An intriguing possibility is that dysregulation of the bile acid metabolism pathway in CD8^+^ TILs is interconnected with tumor microbiome dysbiosis in the TME and may influence the anticancer immune response. It is plausible that manipulating the tumor-associated microbiome may also influence tumor immunity and patient response to immune therapy, as has been demonstrated in the gut microbiome ^42, 50, 51^. Further investigation is needed to explore the interdependence of the tumor microbiome, tumor metabolic reprogramming, and CD8^+^ T cell dysfunction.

Upregulation of bile acid-related genes in tumor infiltrating CD8^+^ T cells correlates with T cell dysfunction activity and may also be related to the differential regulation of metabolic-related pathways such as mTORC1. Connections between bile acid and mTOR pathway activation have been reported. Yamada et al. showed that secondary bile acids activate the mTOR pathway via FXR signaling ^52^. The bile acid activation of mTOR signaling may be caused by upregulation of DGKH (diacylglyerol kinase), an enzyme that converts diacylglycerol to phosphatidic acid which directly activates mTOR ^53^. DGKH was significantly upregulated in tumor infiltrating CD8^+^ T cells in melanoma and SCC in our study.

Our flow cytometric UMAP analysis (Fig. 2) revealed clusters representative of different T cell populations. Dimension reduction of our multiparametric flow cytometry data from peripheral CD8 T cells and CD8^+^ TILs clearly demonstrated that the frequency of cells belonging to cluster 6, with the highest coordinate normalized expression of all immune checkpoint receptors (ICR: CD38, TIGIT, PD-1, TIM-3, and LAG3) as well as CD8 and CD27, was significantly correlated with enrichment of genes from the bile acid metabolism, peroxisome, and fatty acid metabolism pathways in a linear regression model (Fig. 2C). Peroxisome, fatty acid metabolism and glycolysis pathways were also positively correlated with the frequency of cells in cluster 16. We were intrigued to discover that the frequency of cells in clusters that expressed the highest levels of coordinate immune checkpoint receptor expression were significantly positively correlated with enrichment of key metabolic pathways. We propose that TIL clusters 15, 16 and 6 are enriched for CD8^+^ Tex/act activity and may identify progressive levels of immune exhaustion/dysfunction in the CD8 compartment, with progressive coordinate expression of ICR to maximal levels in cluster 6, whose frequency has significant correlation with enrichment of the bile acid metabolism pathway.

Our results are consistent with the hypothesis that transcriptional reprogramming is likely to play a role in driving the changes of cellular metabolism in dysfunctional CD8^+^ TIL in melanoma. TOX, a transcription factor that drives CD8 T cells toward an exhausted state, is upregulated in SCC and melanoma TIL when compared to pCD8 (Fig. 1B and 3A) ^54^, indicating that targeting TOX along with PD-1 could off-set T cell exhaustion. Two key metabolic genes, PKM2 and PHGDH, are upregulated in melanoma TIL compared to healthy pCD8 (3A, and 3E). PKM2 (pyruvate kinase M1/2) expression, which controls glycolysis, has been associated with tumor progression and poor survival in a small pilot study of metastatic melanoma patients ^55^. In addition, PHGDH (D-3-phosphoglycerate dehydrogenase) is overexpressed in multiple cancers, correlates with tumor growth, and has been shown to be increased in 40% of melanoma samples ^56, 57^.

In a subset of our patients that received anti-PD-1 immunotherapy (Fig. 3F), we observed upregulation of CXCR6 and CTTNBP2NL and downregulation of CD160 and HRH2. CXCR6, C-X-C chemokine receptor type 6, is known to be preferentially expressed on CD8^+^PD-1^high^ exhausted T cells in hepatocellular carcinoma (HCC) and is hypothesized to play a role in recruiting CD8^+^ T cells into the tumor microenvironment ^58^. Overexpression of CTTNBP2NL and subsequent STRIPAK complexes could fuel the exhausted CD8^+^ T cells, thus suppressing an immunotherapy effect and promoting tumor progression. These overexpressed refractory gene profiles open new avenues for novel immunotherapy targets. CD160, a degranulation marker, was downregulated in our immune failure signature. Its expression has been demonstrated to be downregulated with repeated antigen stimulation while PD-1 and LAG3 remained high and TIM-3 increased further in T cells from a murine breast cancer model ^59^. Furthermore, in CD8^+^ T cells from individuals with HIV, CD160 and PD-1 co-expression represented a subset of exhausted T cells both functionally and transcriptionally ^60^. Therefore, downregulation of CD160 may indicate dysfunctional TIL that are either no longer degranulating or lack continued TCR engagement. Finally, downregulation of HRH2 (histamine receptor H2) could be indicative of tumor metastasis in the context of immune failure as a balance of histamine with its receptor is necessary to either stimulate or suppress the growth of melanoma ^61^.

scRNASeq and UMAP analysis indicated that cluster 8 correlates with increased mTOR-related signaling, and we identified five genes unique to this cluster (Figure 4C-E). PHGDH, validated in our western blot analysis and scRNAseq, is involved in diverting glycolytic fluxes and is highly expressed in tumors. One study demonstrated it to be amplified at a 40% frequency in melanoma samples ^57^ as reviewed in ^56^. CDK2, E2F1, and BUB1 (Mitotic Checkpoint Serine/Threonine-Protein Kinase BUB1) are involved in cell cycle progression and targeting them could prove to be a viable therapeutic strategy. Inhibition of CDK2 in melanoma cell lines was shown to overcome their resistance to BRAF and Hsp90 inhibitors ^62^ while BUB1 has been identified as a novel target downstream of SIRT1 in melanoma ^63^. E2F1 is overexpressed in melanoma and its inhibition initiates cell cycle arrest/apoptosis as well as increasing the sensitivity of the melanoma cells to BRAF inhibitors ^64^. Finally, PSMA4 was upregulated in OT1 CD8^+^ T cells following transient and continuous Ag-independent stimulation ^65^.

Our highly focused transcriptomic profiling of dysfunctional CD8^+^PD-1^+^TIM-3^+^ TIL in human melanoma and SCC has identified several seemingly disparate and independent metabolic pathways that are differentially regulated in CD8^+^ Tex/act cells in the tumor microenvironment. Singer et al. applied a similar systems immunology approach to analyzing TIL from WT versus MT-/- pMEL mice implanted with B16 murine melanoma cells ^22^. While their study was performed in mice, Singer et al. identified a gene expression signature distinguishing CD8^+^ T cell dysfunction from T cell activation that could also be distinguished (via meta-analysis) amongst a previous scRNAseq study of TIL from human metastatic melanoma tumors ^66^.

We recently employed a biased single cell transcriptomic approach to link higher levels of oxidative phosphorylation in tumor- and peripheral blood–derived total CD8^+^ T cells with ICI resistance in melanoma patients ^67^. Our current systems biology analysis of human melanoma was conducted in an unbiased fashion to fully interrogate dysfunctional CD8^+^PD-1^+^TIM-3^+^ TIL (sorted) transcriptomic profiles, allowing for identification of novel and unexpected signaling pathways that may reveal enhancement targets in more recent immune checkpoint blockade therapies and be more capable of discerning responders from nonresponders in broad cohorts. Taken together, our findings indicate that mTOR, bile acid metabolism, and peroxisome metabolic pathways are coordinately dysregulated and highly upregulated in the CD8^+^ TIL of melanoma and SCC patients, impacting the capacity of the host immune system to suppress and eradicate tumor cells. The discovery of novel metabolic targets in Tex/act cells could also reinvigorate resistance in immune checkpoint therapy failure, thus improving clinical efficacy. Our results provide a new platform and public resource for datamining and developing novel treatment modalities to further improve current immunotherapy advancement.

## Methods

### Patient selection and demographics

All human tissue was obtained at the Cleveland Clinic under a protocol approved by Cleveland Clinic’s institutional review board, and written informed consent was obtained from each patient. Peripheral blood lymphocytes (PBLs) and tumor specimens were obtained from patients with cutaneous melanoma (MEL, n=23), squamous cell carcinoma (SCC, n=8), and control individuals without skin disease (n=8) as previously described ^18^. MEL patients were 60.9% male with a mean age of 58 (±15.8); 3 patients presented with a primary tumor while 20 patients had metastatic disease. Fourteen of the MEL patients received some type of immunotherapy (IO), out of these patients, 8 received anti-PD-1/PD-L1 directed IO (either pembrolizumab or nivolumab). SCC patients were 85.7% male with a mean age of 73 (±7.9); 3 patients presented with a primary tumor while 5 patients had metastatic disease. Age and sex are unknown for one SCC patient. Tissue for scRNAseq libraries was obtained from patients with cutaneous melanoma (n=8). The patient cohort was 37.5% male with a mean age of 60 (±12.7). 2 patients presented with a primary tumor while 6 patients had metastatic disease.

### Isolation of TILs and PBLs

After surgical resection, tumor specimens were washed with antibiotic-containing media and minced with crossed scalpels under sterile conditions. Tissue was dissociated via enzymatic digestion using 1,500 U/ml collagenase IV (Gibco/Life Technologies), 1,000 U/ml hyaluronidase (Sigma), and 0.05 mU/ml DNase IV (Gibco) in RPMI for 1 h at 37°C followed by mechanical agitation. Centrifugation over Ficoll-Hypaque gradient was used to separate debris from the single cell suspension. Finally, cells were cryopreserved in 10% DMSO + bovine serum. Similarly, peripheral blood mononuclear cells were purified from buffy coats by centrifugation over a Ficoll-Hypaque gradient followed by cryopreservation.

### Flow cytometric analysis

Flow cytometric analysis of immune checkpoint receptor and activation marker expression and T cell memory subset distribution was carried out as follows: for memory subset phenotyping and assessment of negative regulator and ligand expression, a cocktail of the following monoclonal antibodies was used: anti-CD3 Alexa700 (BD), anti-CD4 Q.605 (Invitrogen), anti-CD8 PerCP (Biolegend), anti-CD45RA BV650 (Biolegend), anti-CD27 APC-eFluor 780 (eBioscience), anti-CCR7 PE-CF594 (BD), anti-CD14 V500 (BD), anti-CD19 BV510 (Biolegend), Live/Dead Amcyan (Invitrogen), anti-BTLA (BD), anti-TIM3 BV421 (Biolegend), anti-PD-1 PE-Cy7 (Biolegend), anti-CTLA-4 APC (BD), anti-TIGIT PE (eBioscience), anti-LAG3 FITC (Novus). After washing, cells were resuspended in staining buffer and sorted on an ARIA-SORP. The following antibodies were used for intracellular flow cytometry to assess phosphorylation of mTOR targets: anti-S6 (S235/S236) V450 (BD), anti-p4E-BP1 (T36/46) PE (BD). For the rapamycin assay, cells were stimulated for 3 days with CD3/CD28 beads and then treated with 25nM rapamycin overnight. Cells were stained and analyzed for PD-1 surface expression. The following antibodies were used to identify memory T cells and effector memory T cells: anti-CD8 BV711 (BD), anti-CD45RA BV650 (Biolegend), anti-CCR7 FITC (R&D), anti-CD27 APC eFluor 780 (eBioscience). Data were analyzed using FlowJo software (TreeStar) and our custom pipeline for dimensionality reduction.

### Western blot

Isolated CD8 cells (50,000) from TIL were lysed in 50 uL RIPA buffer (150mM NaCl, 1mM EDTA, 0.5% Triton X-100, 0.5% deoxycholic acid, 0.5% SDS, and 100mM Tris (pH 7.5) containing protease and phosphatase inhibitors. An equal sample volume (100ul) was used for reducing gel electrophoresis (4-12%) followed by immunodetection with antibodies toward CYP27A1 (Abcam), Actin (Abcam), PHGDH (Cell Signaling), PKM2 (Cell Signaling), and PCK2 (Cell Signaling). Three biological replicates were performed.

### Bulk RNA-seq and bioinformatic analysis

RNA was purified from CD8 T cells using RNeasy Micro Kits (Qiagen), followed by low input RNASeq library generation using Takara SMART-Seq v4 Ultra Low/Nextera XT with Nextera Index v2 Set A. Paired-end sequencing reactions were run on an Illumina NextSeq 550 High Output platform (25M total reads per sample). Raw demultiplexed fastq paired-end read files were trimmed of adapters and filtered using the program skewer to remove reads with an average phred quality score of less than 30 or trimmed to a length of less than 36 ^68^. Trimmed reads were then aligned using the HISAT2 aligner to the Homo sapiens NCBI reference genome assembly version GRCh38 and sorted using SAMtools ^69, 70^. Aligned reads were counted and assigned to gene meta-features using the program featureCounts as part of the Subread package ^71^. These count files were imported into the R programming language and were assessed for quality control, normalized and analyzed, utilizing the limma-trend method ^72^ for differential gene expression testing as well as regression modelling and GSVA ^73^. Linear regression modelling was performed using the limma framework. RNA-seq data supporting this study have been deposited in the Gene Expression Omnibus (GEO) public database with the accession number pending.

### scRNAseq Library Preparation and Data Processing

All cells were resuspended in DPBS with 0.04% BSA and immediately processed for scRNAseq as follows. Cell count and viability were determined using trypan blue on a Countess FL II, and approximately 12,000 cells were loaded for capture onto the Chromium System using the v2 single cell reagent kit according to the manufacturer’s protocol (10X Genomics). Following capture and lysis, cDNA was synthesized and amplified (12 cycles) as per manufacturer’s protocol (10X Genomics). The amplified cDNA from each channel of the Chromium System was used to construct an Illumina sequencing library and was sequenced on an Illumina HiSeq 2500 with 150-cycle sequencing (asymmetric reads per 10X Genomics). Illumina basecall files (*.bcl) were converted to FASTQs using CellRanger v3.0, which uses bcl2fastq v2.17.1.14. FASTQ files were then aligned to GRCh38 human reference genome and transcriptome using the CellRanger v3.0 software pipeline with default parameters as reported previously ^74^; this demultiplexes the samples and generates a gene versus cell expression matrix based on the barcodes and assigns UMIs that enables determination of the individual cell from which the RNA molecule originated. Overall, an estimated total of 71,238 cells were analyzed.

### scRNASseq Bioinformatic Analysis

Bioinformatic analysis of cells based on whole transcriptomes was performed by the R package Seurat (version 3.0) ^75^. Dimensionality of gene-barcode matrices was first reduced to 18 principal components using principal components analysis (PCA). PCA-reduced data were further reduced to 2-dimensional space using the UMAP method and visualized. Graph-based clustering of cells was conducted in the PCA space; a sparse nearest-neighbor graph of the cells was built first and Louvain modularity optimization was then applied. The number of nearest neighbors was logarithmic in accordance with the number of cells. In the last step, repeated cycles of hierarchical clustering and merging of cluster pairs that had no significant differential expression were performed, until no more cluster pairs could merge. Differential gene expression analyses of each cluster were conducted by Wilcoxon rank sum test. The log2 fold-change of a certain gene’s expression (UMIs) in one cluster vs. all other clusters, and the corresponding adjusted p-values, were calculated for each cluster with pathway enrichment scores generated using the Gene Set Enrichment Analysis (GSEA) method ^76^.

### Statistics

Unless otherwise indicated, the Student’s t-test was used with p≤0.05 chosen as the level of significance and the analysis was performed with Graphpad Prism 8.

## Data Availability

RNA-seq data supporting this study have been deposited in the Gene Expression Omnibus (GEO) public database with accession number pending.

## Abbreviations

Tex/act: exhausted but activated T cell subpopulation
TIL: tumor infiltrating lymphocyte
mTOR: mammalian target of rapamycin
ICR: immune checkpoint receptor
TME: tumor microenvironment
PD-1: programmed cell death-1
SCC: cutaneous squamous cell carcinoma
pCD8: peripheral CD8 T cells
MEL: melanoma
IO: immunotherapy
RIPA: radioimmunoprecipitation assay buffer
GEO: Gene Expression Omnibus
PCA: principal components analysis
UMAP: uniform manifold approximation and projection
MFI: mean fluorescence intensity
HCC: hepatocellular carcinoma;
GSVA: Gene Set Variation Analysis
DEG: differentially expressed genes
PBMC: peripheral blood mononuclear cells
scRNAseq: single cell RNA sequencing
Tim-3: T cell immunoglobulin and mucin protein 3

## Acknowledgments

We thank the Genomics Core at the Lerner Research Institute of Cleveland Clinic and the Genomics and Applied Functional Genomics Cores at Case Western Reserve University for their technical and analytical support. This research is supported by Cleveland Clinic and Case Western Reserve University internal funding.

---

**Supplemental Figure 1:**
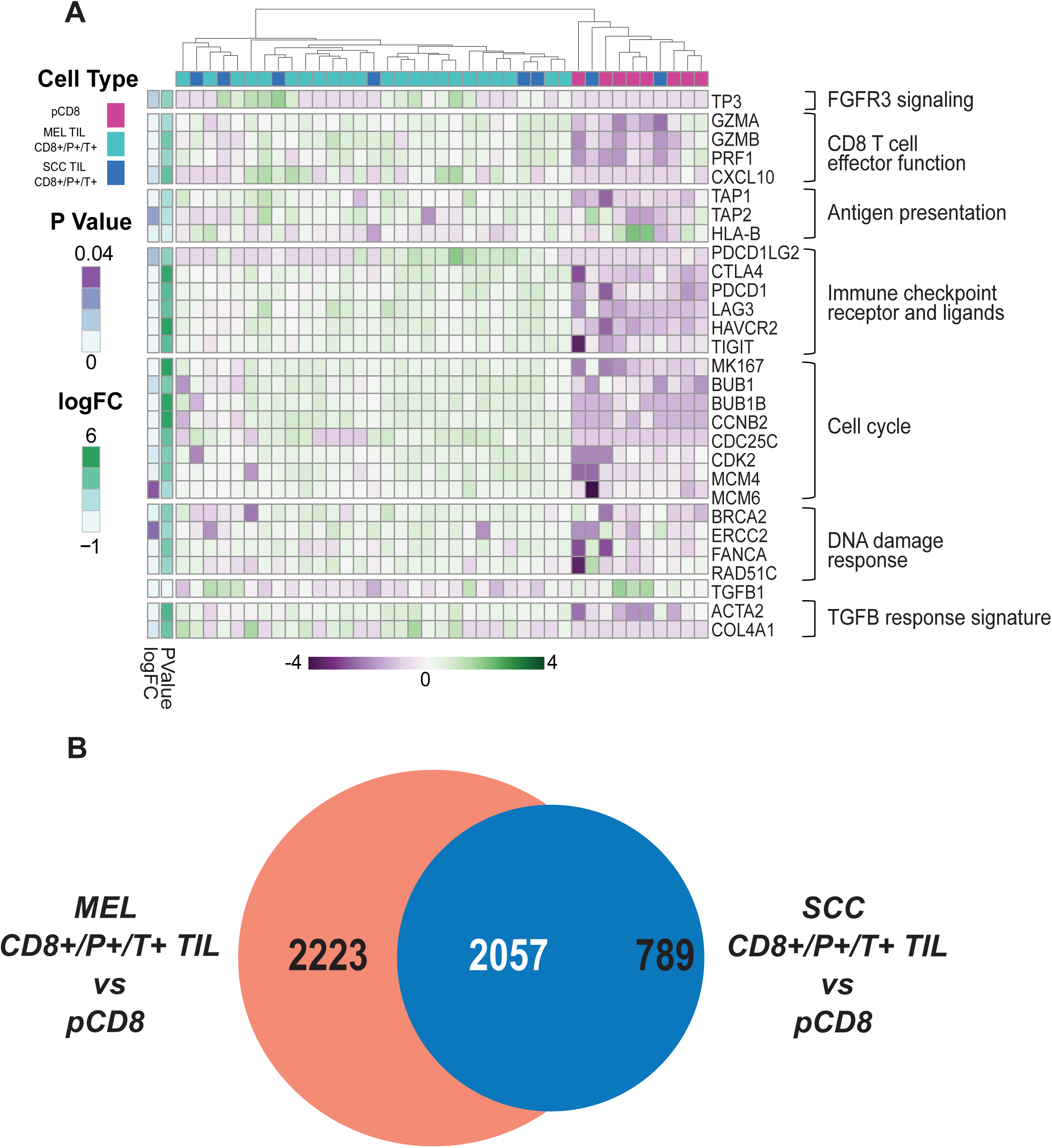
Characterization of dysfunctional CD8+ T cells in total TIL and PBMC. **(A)** Venn analysis identified genes that are common and unique in total (melanoma and SCC) CD8^+^PD-1^+^TIM-3^+^ TIL (nominal p≤0.05). 2,057 genes were commonly differentially regulated in both total CD8^+/^P^+/^T^+^ TIL and peripheral CD8^+^ T cells, while 2,223 and 789 were unique to melanoma and SCC CD8^+/^P^+/^T^+^ TIL, respectively, in comparison to healthy peripheral CD8^+^ T cells. **(B)** Core biological pathways associated with TGF-β attenuation of the tumor response to immune checkpoint blockade were significantly differentially regulated within the dysfunctional CD8^+/^P^+/^T^+^ melanoma and SCC TIL as shown by the two way heatmap. Pathways and genes (noted on the right y-axis) include those involved in FGFR3 signaling (TP63), CD8 T cell effector function (GZMA, GZMB, PRF1 and CXCL10), immune checkpoint receptors and their ligands (PDCD1LG2, CTLA4, PD-1, LAG3, TIM-3, and TIGIT), cell cycle (MKI67, CCNE1, BUB1, BUB1B, CCNB2, CDC25C, CDK2, MCM4, and MCM6), DNA damage response (BRCA2, ERCC2, FANCA, and RAD51c), and TGF-β response signature genes (ACTA2 and COL4A1). HLA-B and TGFB1 genes were uniquely downregulated in the CD8^+^ TIL as compared to PBMC. All p≤0.05.

**Supplemental Figure 2:**
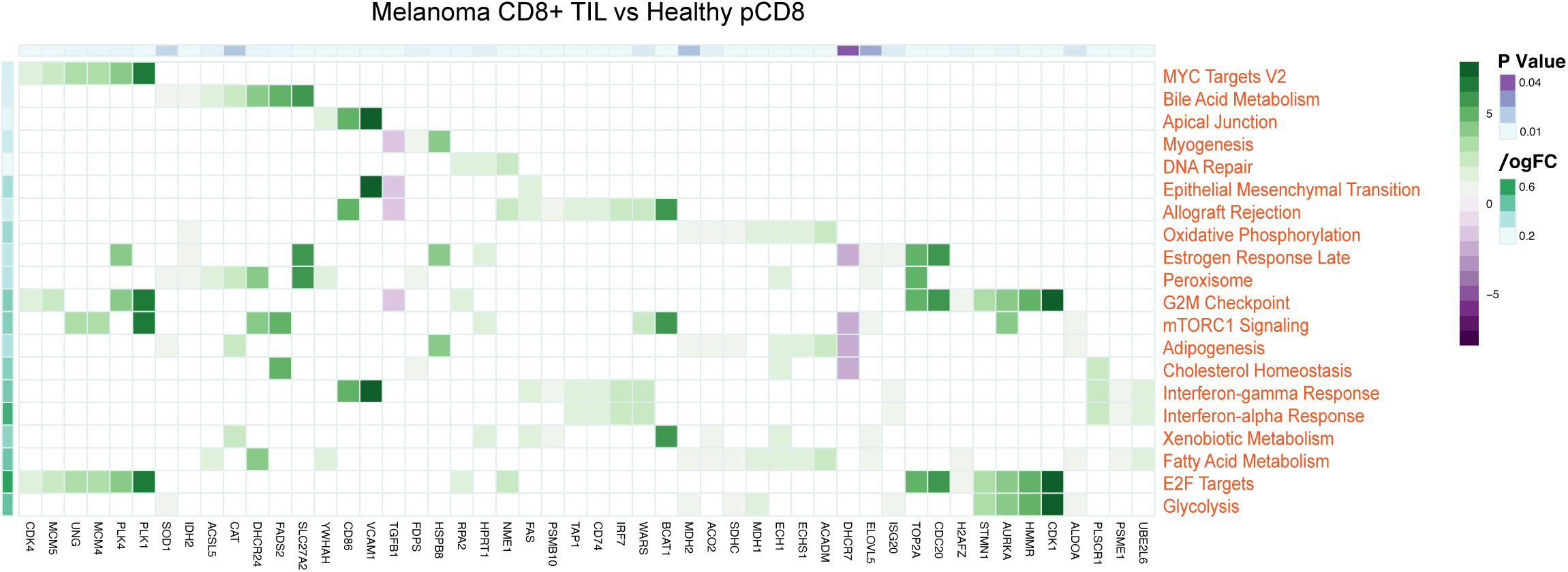
Hallmark pathway enrichment in melanoma TIL. Top 20 differentially enriched pathways between melanoma TILs and pCD8s with associated top 50 shared differentially expressed genes. Pathways and genes were selected based on a nominal p≤0.05 and the number of genes in common between pathways. Heatmap values show the logFC values for each gene between melanoma TILs and pCD8s while their column annotations denote p values. Row annotations denote the logFC values of pathway enrichment scores between the two groups.

